# Multi-task deep learning integrating pretreatment MRI and whole slide images predicts induction chemotherapy response and survival in locally advanced nasopharyngeal carcinoma

**DOI:** 10.64898/2026.04.07.26350350

**Authors:** Jing Hou, Xiaochun Yi, Congrui Li, Junjian Li, Hui Cao, Qiang Lu, Xiaoping Yu

**Author notes:** Corresponding Author (XY).

## Abstract

Predicting response to induction chemotherapy (IC) and overall survival (OS) is critical for optimizing treatment in patients with locally advanced nasopharyngeal carcinoma (LANPC). This study aimed to develop and validate a multi-task deep learning model integrating pretreatment MRI and whole slide images (WSIs) to predict IC response and OS in LANPC. Pretreatment MRI and WSIs from 404 patients with LANPC were retrospectively collected to construct a multi-task model (MoEMIL) for the simultaneous prediction of early IC response and OS. MoEMIL employed multi-instance learning to process WSIs, PyRadiomics and a convolutional neural network (ResNet50) to extract MRI features, and fused multimodal features through a multi-gate mixture-of-experts architecture. Clustering-constrained attention multiple instance learning and gradient-weighted class activation mapping were applied for visualization and interpretation. MoEMIL effectively stratified patients into good and poor IC response groups, achieving areas under the curve of 0.917, 0.869, and 0.801 in the train, validation, and test sets, respectively, and outperformed the deep learning radiomics model, the pathomics model and TNM staging. The model also stratified patients into high- and low-risk OS groups (*P < 0.05*). MoEMIL shows promise as a decision-support tool for early IC response prediction and prognostication in LANPC.

**Author Summary:** We have developed a deep learning model that integrates two types of medical images, including magnetic resonance imaging (MRI) and digital pathological slices, to simultaneously predict response to induction chemotherapy and prognosis in patients with locally advanced nasopharyngeal carcinoma. Current treatment decisions primarily rely on traditional tumor staging (TNM), which often fails to comprehensively reflect the complexity of the disease. Our model, named MoEMIL, was trained and tested on data from 404 patients across two hospitals and consistently outperformed both single-model approaches and TNM staging methods. By identifying patients who exhibit poor response to induction chemotherapy or higher prognostic risk, our tool can assist clinicians in achieving personalized treatment, enabling intensified management for high-risk patients and avoiding unnecessary side effects for low-risk patients. Additionally, we visualize the model’s reasoning process through heat map generation, which highlights the image regions exerting the greatest influence on prediction outcomes. This work represents a step toward more precise treatment for nasopharyngeal carcinoma; however, larger-scale prospective studies are required before the model can be integrated into routine clinical practice.

## Introduction

Nasopharyngeal carcinoma (NPC) is an epithelial malignancy with a distinct geographical prevalence in East and Southeast Asia[1,2]. Approximately 70–80% of patients are diagnosed with locally advanced nasopharyngeal carcinoma (LANPC), which is generally classified as stage III or IVa[1]. The current standard treatment for LANPC involves induction chemotherapy (IC) followed by concurrent chemoradiotherapy (CCRT)[3], which has been shown to improve survival outcomes compared to CCRT alone. However, not all patients respond well to IC. Moreover, approximately 20%-30% of LANPC patients ultimately experience treatment failure[4]. Local recurrence and distant metastasis are the main reasons for treatment failure. Tumor response to IC is an independent prognostic factor for survival after intensity-modulated radiation therapy in NPC[5]. Accurate risk assessment following IC could guide timely adjustments to subsequent CCRT, potentially improving survival while reducing unnecessary treatment-related toxicity.

The Tumor-Node-Metastasis (TNM) staging system remains the primary method for guiding therapeutic decisions and prognostication in patients with LANPC[6]. However, its reliance on anatomical details overlooks the inherent heterogeneity within tumors, often leading clinicians to observe disparities in early treatment response and overall prognosis among patients staged identically according to TNM. This limitation underscores the urgent need for more refined approaches that can capture the complex biological characteristics of tumors.

Medical imaging provides a noninvasive way to monitor tumor response and progression during treatment. Advances in radiomics enable the transformation of visual medical images into mineable high-dimensional data through high-throughput computing, a technique that has been widely applied to predict tumor response and survival outcomes in the early stage of LANPC. Compared with conventional radiomics, deep learning automatically quantifies complex tumor characteristics beyond human visual perception and has attracted considerable attention in medical imaging research due to its superior performance. Deep survival models have demonstrated the potential to outperform conventional radiomics-based prognostic prediction models[7]. However, most current models for IC prediction and prognosis focused on performing a single task method[8,9], unable to capture inter-task relationships or effectively model the complex nonlinear interactions between various prognostic factors[8,10]. In contrast, multi-task learning allows multiple related tasks to be learned simultaneously within a single model. By sharing feature representations between related tasks, multitask learning is more data-efficient and has been shown to reduce overfitting and improve model generalizability in many tumors, including gastric cancer[11,12], lung cancer[13].

In addition to radiological imaging, pathological examination plays a crucial role in diagnosing NPC, directing clinical treatment, and forecasting prognosis. The former offers macroscopic details about the tumor, whereas the whole-slide image (WSI) of the latter can thoroughly disclose the biological traits and microenvironment of the tumor. The integration of radiological and pathological data, known as radiopathomics, has shown promise in enhancing predictive accuracy by capturing tumor information at different scales. This approach has been successfully applied to prognosis prediction and therapeutic efficacy assessment in various solid tumors, including breast cancer[14], prostate cancer[15], and rectal cancer[16]. However, its application in NPC remains limited, with few studies leveraging the complementary information provided by these multimodal data sources, especially in the multi-task prediction.

In light of these considerations, we aimed to develop and validate an integrative radiopathomics model based on deep learning for simultaneously predicting response to IC and clinical outcomes in patients with LANPC. The model employs multi-instance learning (MIL) to process WSIs, a convolutional neural network (ResNet50) to extract MRI features, and integrates these multimodal features through a multi-gate mixture of experts (MMoE) architecture[17]. By leveraging multi-center data and explainable artificial intelligence techniques, we sought to create a robust, interpretable predictive tool that can ultimately support personalized treatment planning for LANPC patients, addressing the limitations of current unimodal and single-task approaches.

The main aims of this study were (1) the development of a novel multi-task deep learning framework that simultaneously predicts IC response and overall survival (OS) in patients with LANPC; (2) the integration of MRI and WSI data to capture complementary information at different scales; and (3) comprehensive validation using multi-center data and demonstration of clinical utility through survival stratification.

## Materials and Methods

### Patients

This retrospective study was approved by the institutional review board, and the requirement for written informed consent was waived. A total of 404 eligible LANPC patients were included in this study. The primary cohort consisted of 364 patients retrospectively enrolled from Hunan Cancer Hospital (January 2014 to June 2018). An external test cohort of 40 patients was recruited from the First Affiliated Hospital of Jinan University (Guangzhou, China). Inclusion criteria were: (1) histologically confirmed NPC, classified as stage III to IVa according to the 7th before 2017 or 8th after 2017 edition of the AJCC TNM staging system; (2) received IC followed by CCRT; (3) availability of complete pretreatment contrast-enhanced T1-weighted (CET1-w) MRI sequences; and (4) availability of high-quality hematoxylin and eosin (H&E)-stained biopsy slides obtained via nasopharyngoscopy. Exclusion criteria were: (1) history of other malignancies or prior anticancer therapies; (2) poor-quality H&E slides (e.g., excessive staining artifacts); and (3) incomplete efficacy assessment data after IC and follow-up data. The patient selection process is detailed in Fig 1.

**Fig 1.**
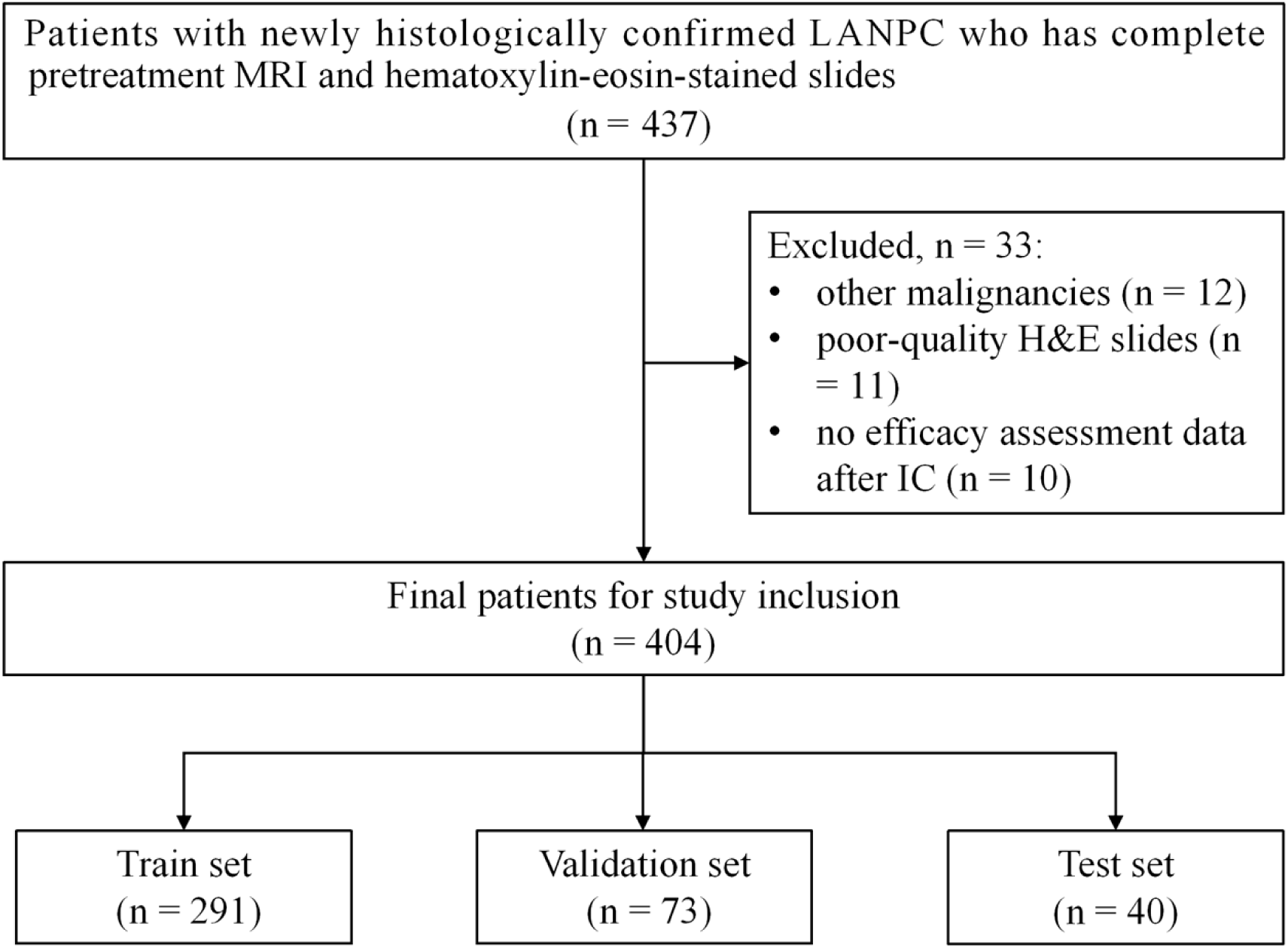
Flow-chart shows patient recruitment process. Abbreviations: LANPC = local advanced nasopharyngeal carcinoma, MRI = magnetic resonance imaging, IC = induction chemotherapy

IC regimens were cisplatin-based, administered every 3 weeks for 2-4 cycles. Radiotherapy was delivered with a total dose of 70-76 Gy in 30-33 fractions. Concurrent chemotherapy during radiotherapy involved cisplatin for 3-6 cycles. Follow-up was conducted per National Comprehensive Cancer Network (NCCN) guidelines[3]. Nasopharyngeal MRI follow-up evaluations were performed every 3 months in the first year, every 6 months in the second year, and annually thereafter. The curative response to IC was classified into complete response(CR; with no evidence of residual disease on MRI and nasopharyngoscopy, regression ratio of 100%), partial response (PR; with evidence of residual disease, regression ratio >30%), stable disease (SD; with evidence of residual disease, regression ratio <30%) and progressive disease (PD; with evidence of increase, increase ratio>20%) according to the Response Evaluation Criteria in Solid Tumors (RECIST) 1.1 guidelines[18]. CR and PR are considered as good responders, while SD and PD are classified as poor responders. The primary endpoint was OS, defined as the time from diagnosis to death from any cause.

### MRI acquisition and segmentation

MRI scans were conducted using a 1.5-Tesla MRI scanner with a combined head and neck coil. The MR acquisition protocols are presented. At our hospital, a 1.5-Tesla MRI system equipped with a combined head-and-neck coil was utilized to acquire the scans. The imaging sequence parameters were: Axial T1-weighted spin-echo: repetition time (TR)/echo time (TE) = 580/7.8 ms, slice thickness = 5 mm, 36 slices, 1 mm interslice spacing, and number of excitations (NEX) = 2. Axial fat-suppressed T2-weighted spin-echo: TR/TE = 6289/85 ms, slice thickness = 5 mm, 36 slices, 1 mm spacing, NEX = 2. Axial contrast-enhanced T1-weighted spin-echo (CET1-w): TR/TE = 500/8 ms, field of view (FOV) = 22 × 22 cm, NEX = 2, slice thickness = 4 mm, interslice gap = 0.8 mm. For the CET1-w images, a gadolinium-based contrast agent (gadodiamide) was injected intravenously at a dose of 0.1 mmol/kg body weight, with a flow rate of 3.5 ml/s. At the First Affiliated Hospital of Jinan University, MRI acquisitions were performed on another 1.5 T scanner. The protocols were: Axial T1-weighted imaging (T1WI): TR/TE = 500/8 ms, slice thickness = 4 mm, interslice spacing = 1 mm, NEX = 2. Axial T2-weighted imaging (T2WI): TR/TE = 5000/85 ms, slice thickness = 4 mm, spacing = 1 mm, NEX = 2. Axial CET1-w: TR/TE = 500/8 ms, FOV = 22 × 22 cm, slice thickness = 4 mm, spacing = 1 mm, NEX = 2.

MRI preprocessing was conducted using AK software (Artificial Intelligence Kit, GE Healthcare), which has been officially registered and approved. The preprocessing involved resampling, skull stripping, and intensity normalization. The image resolution was adjusted to 1 × 1 × 1 mm during resampling, with all MRI image thicknesses standardized to 1 mm through linear interpolation. A gray-level adjustment was then applied to standardize the intensity range to 0–255. Two experienced radiologists (with 16 and 15 years of experience in head and neck MRI, respectively), blinded to clinical outcomes, independently delineated the primary tumor volume of interest (VOI) on each MRI scan using ITK-SNAP software (version 3.8.0; http://www.itksnap.org/).

### Radiomics features and deep learning features extraction

To capture comprehensive tumor phenotypic information, we designed a dual-branch feature extraction framework that integrates traditional radiomics features with deep learning-derived features. Radiomics features were extracted using the IBSI-compliant PyRadiomics toolkit to quantify tumor texture, shape, and intensity characteristics[19]. A total of 107 features were computed, including 14 morphological descriptors, 18 first-order statistical features, and 75 second-order texture features derived from gray-level co-occurrence matrices (GLCM), gray-level run-length matrices (GLRLM), gray-level size zone matrices (GLSZM), neighborhood gray-level difference matrices (NGTDM), and gray-level dependence matrices (GLDM). To construct a unified feature set, we treated the distinct radiomics categories (morphology, statistics, texture) and the deep features as radiology-based instances and fed them into instance-wise fully connected layers, respectively, to generate radiology embeddings *R_bag_*= {r_l_}∈R^L×dr^. In parallel, the same VOIs were fed into a pre-trained ResNet-50 convolutional neural network (CNN) with its final classification layer removed to extract deep learning features. The resulting deep features were also transformed through a separate linear layer, to generate CNN embeddings *R_deep_*∈R^l×dr^. Finally, the radiomics features and deep CNN features were fused using an attention mechanism to form a unified radiological feature representation for downstream analysis.

### Histological feature encoding from WSIs

Histological feature extraction was performed using a deep learning-based, weakly supervised framework adapted from Clustering-constrained Attention Multiple Instance Learning (CLAM)[20]. H&E-stained tissue sections were digitally scanned at 40× magnification using a Pannoramic MIDI II scanner, yielding WSIs in SVS format with a resolution of 0.14 µm/pixel. Following the CLAM preprocessing pipeline, Otsu’s thresholding was first applied to separate foreground tissue from the background. Given the gigapixel size of WSIs, which prevents direct network input, the tissue regions were segmented into non-overlapping patches of 448 × 448 pixels. Subsequently, each patch was encoded using the pre-trained CONCHv1.5 visual encoder, which serves as the patch-level feature extractor of the TITAN foundation model, to obtain high-quality latent representations[21]. To aggregate these patch features into a single slide-level representation, an attention-based Multiple Instance Learning (MIL) framework was employed[22,23]. In this approach, all patches from a WSI are treated as a “bag” of instances. The network learns attention weights that reflect the diagnostic relevance of each patch, and this is followed by a weighted summation of the patch features. This process allows the model to focus on morphologically informative regions while suppressing contributions from non-tissue background, ultimately generating a compact, global feature vector for the entire WSI. Furthermore, attention heatmaps were generated based on the attention scores derived from the pooling layer to visualize the importance of each region within the WSI.

### Multimodal Feature Fusion via MMoE

Before the Multi-gate Mixture-of-Experts (MMoE) module, the above-extracted radiological and pathological feature vectors were initially fused and concatenated to form a combined multimodal representation. Subsequently, this combined feature vector was sent to a fusion module, which consists of a linear layer for dimensionality reduction and feature mapping, followed by the ReLU activation function and Dropout regularization to introduce nonlinearity and reduce overfitting. This step not only standardizes the feature dimensions but also prepares the fused representation for processing in the subsequent MMoE module.

The MMoE module serves as the core innovative component of the model, designed to enable effective multi-task feature sharing. This process consists of three parts, namely the expert network, the screening network, and the weighted feature fusion. Expert Network: This module utilizes multiple feed-forward neural networks (i.e., experts), which are structurally identical but have independent parameters. During the implementation process, each expert is implemented as a two-layer multi-layer perceptron (MLP), with a structure of linear → ReLU → Dropout → linear → ReLU. All specific task features from the previous fusion stage are processed in parallel through these expert networks, thereby generating a set of diverse feature representations. Gated Network: Two independent gate-controlled networks are respectively set up for two different downstream tasks, namely IC response evaluation and survival prediction. Each gate-controlled network acts as a lightweight attention mechanism, which receives input features and outputs a normalized weight vector (for example, [0.2, 0.1, 0.7]), reflecting the relevance of each expert’s output to the current task. Weighted Feature Fusion: For each specific task, the weighted vector calculated by the corresponding gating network is multiplied to the outputs of each expert network and then summed. Through this dynamic weighting mechanism, each task ultimately obtains a feature representation tailored to it, which is composed of different expert features according to the task requirements, thereby effectively alleviating the problem of negative transfer.

### MoEMIL model construction and validation

Following the MMoE module, this component generates adaptive feature representations tailored to the specific task. These features are then routed to distinct prediction heads to produce the final clinical output. The IC response evaluation head receives the first task-specific feature vector from the MMoE output and processes it through a Multi-Layer Perceptron (MLP). The MLP output corresponds to classification prediction values for different response categories. The survival prediction head processes the second task-specific feature vector via another MLP to generate a risk score. This score can be utilized to generate Kaplan-Meier survival curves or calculate Cox proportional hazards loss, thereby facilitating the assessment of patient prognosis.

For the IC response prediction task, we employed the cross-entropy loss function (*L_IC_*), while for the prognosis prediction task (OS), the discrete-time negative log-likelihood loss function (*L_OS_*) was utilized. The total loss was defined as the sum of these two components: *L_total_* = *L_IC_* + *L_OS_*. All models were trained for a maximum of 200 epochs with a batch size of 1 and an initial learning rate of 2 × 10^-4^. To prevent overfitting, an early stopping strategy was implemented with a patience of 8 epochs. All model training and inference processes were performed on an NVIDIA GeForce RTX 3080 Ti GPU.

The predictive and prognostic performances of the Deep Learning Radiomics (DLR), Pathomics, and integrated model named Mixture-of-Experts integrated Multiple Instance Learning (MoEMIL) models were quantitatively evaluated using the receiver operating characteristic (ROC) curve and concordance index (C-index). Additionally, Kaplan–Meier curve analysis was conducted to demonstrate the models’ ability to stratify LANPC patients into high- and low-risk groups. The workflow of the study is depicted in Fig 2.

**Fig 2.**
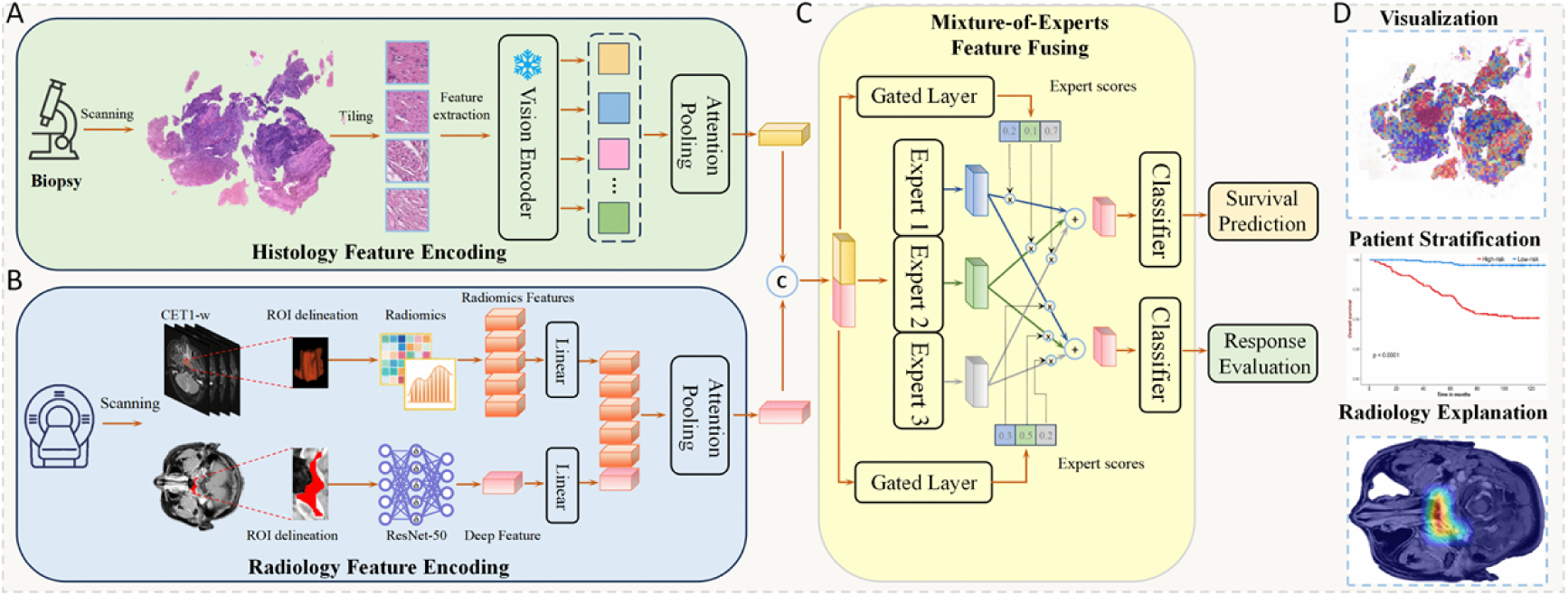
Workflow of this study. The entire model architecture consists of three core components, including the feature encoding module (MRI and WSI feature encoding), the multimodal fusion module, and the task-specific output heads. (A) Histology feature encoding: WSIs are processed by first cropping them into clinical-grade patches of 448 × 448 pixels at 20× magnification. These patches are then fed into the CONCHv1.5 visual encoder (from the TITAN foundation model) to extract high-dimensional latent representations. An attention pooling mechanism is subsequently employed to aggregate these patch-level features into a global histological feature vector, effectively identifying the most diagnostically relevant regions. (B) Radiology feature encoding: This module extracts complementary information from multi-parametric MRI (including CET1-w sequences). It combines hand-crafted radiomic features (capturing texture and shape) with deep CNN features (extracted via a ResNet-50 backbone). These heterogeneous radiological signatures are fused through an attention-based bottleneck to generate a unified radiology feature representation. (C) Mixture-of-experts feature fusion: This process utilizes an MMoE architecture. The fused multimodal input is processed by a set of shared experts, while dedicated gated layers learn to assign specific weights (expert scores) to each expert’s output. This allows the model to dynamically balance the contribution of histological and radiological data based on the specific requirements of each downstream task. (D) Model visualization and clinical application: For WSIs, attention heatmaps highlight morphological regions contributing to high-risk scores. For MRI, Grad-CAM generates saliency maps to visualize the volumetric basis of the model’s decision-making. Abbreviations: CET1-w = contrast-enhanced T1-weighted spin-echo image, WSI = whole slide image, MIL = multiple instance learning, Grad-CAM = Gradient-weighted Class Activation Mapping

### Model interpretation and visualization

To interpret the importance of different tissue regions for prognostic predictions in a WSI, attention scores were computed for all non-overlapping 256×256 tissue patches within a WSI. These scores were normalized to a percentile scale from 0.0 (low attention) to 1.0 (high attention) and spatially registered back to the original slide. For visualization, the scores were converted into an RGB heatmap using a color map and overlaid on the corresponding H&E image at 50% transparency. Regions with high attention (typically colored red) frequently corresponded to morphologically distinct tumor areas, whereas regions with lower attention (often blue) included normal tissue and background. This method provides an intuitive, annotation-free visualization of the tissue regions that the model deems most relevant for prognosis.

For interpretation and visualization of MRI, gradient-weighted class activation mapping (Grad-CAM) algorithm was used to identify and visualize the regions of the MRIs that play a crucial role in the model ’ s predictions[24]. This method provides explainability by visualizing, in the form of a heatmap, the area on which the model focuses when making predictions, intuitively showing the basis used by the model to make decisions. The code and model weights are publicly available at https://github.com/junjianli106/PathRadMoE.

### Statistical analysis

Quantitative data were presented as mean (standard deviation [SD]) or median (interquartile range [IQR]). Continuous variables were compared using Student’s t test or Wilcoxon signed-rank tests, and categorical variables were compared using the χ² test or Fisher’s exact test. All statistical analysis was two-sided, and *p* values of less than 0.05 indicated statistical significance. Area under the curve (AUC) of the receiver operating characteristic (ROC) curve was applied to assess the accuracy of the DLR, pathomics, and MoEMIL models for predicting IC response. Kaplan–Meier curve and log-rank tests were applied to assess survival outcomes among subgroups. Concordance index(C-index) was used to assess the prognostic performance of the DLR, pathomics, and MoEMIL models. Data processing and analysis were performed using R version 4.3.3 (2024-02-29), along with Zstats 1.0 (www.zstats.net). All statistical analysis were performed using R version 4.3.0 (2023-04-21), along with Storm Statistical Platform (www.medsta.cn/software), or Python (version 3.8.18) with the scikit-learn package (version 1.3.2), PyTorch (version 2.1.2), NumPy (version 1.24.1), Pandas (version 2.0.3), and OpenSlide (version 1.3.1).

## Results

### Patient characteristics

The initial sample group of this study consisted of 474 patients with LANPC. Among them, 33 patients were excluded due to having other malignant tumors (n = 12), poor quality of H&E slides (n = 11), and no efficacy assessment data after IC (n = 10). Finally, 404 LANPC patients were included in the study (265 men, 139 women), with a mean age of 54.00 ± 7.07 years. These patients were allocated into a train set (n = 291), a validation set (n = 73), and a test set (n = 40). The median follow-up duration was 87.63 months (interquartile range, IQR: 53.00-109.03). Baseline clinical characteristics, including age, sex, T stage, N stage, TNM stage, and differentiation degree, showed no significant differences across the three cohorts (Table 1), which confirmed a well-balanced data partition. The available EBV-DNA data were insufficient to perform a meaningful statistical analysis on these three datasets.

**Table 1.**
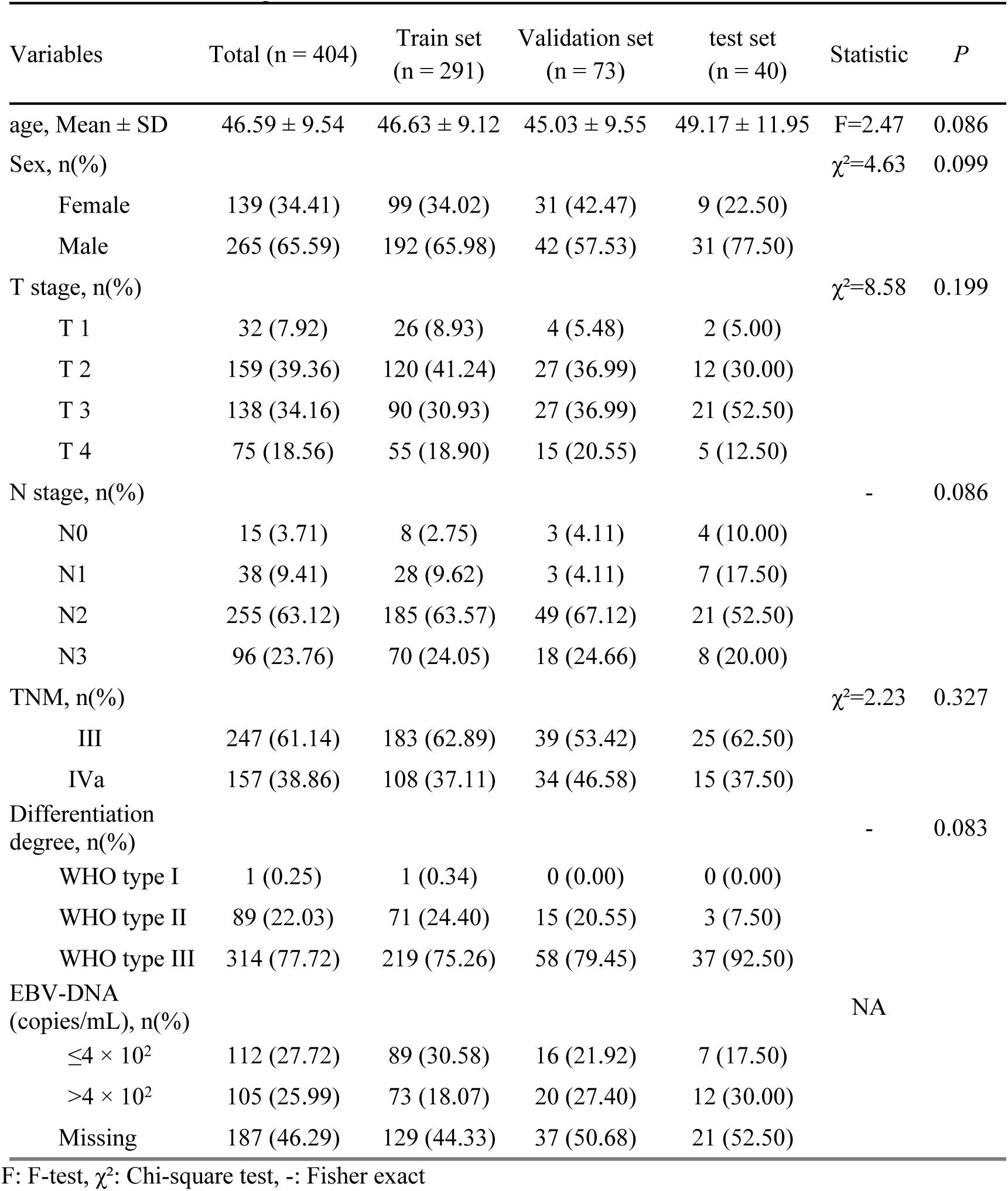
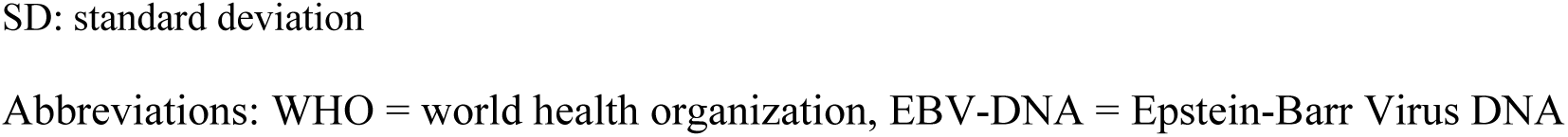
Characteristics of patients in the train, validation, and test sets.

| Variables | Total (n = 404) | Train set<br>(n = 291) | Validation set<br>(n = 73) | test set<br>(n = 40) | Statistic | P |
| --- | --- | --- | --- | --- | --- | --- |
| age, Mean ± SD | 46.59 ± 9.54 | 46.63 ± 9.12 | 45.03 ± 9.55 | 49.17 ± 11.95 | F=2.47 | 0.086 |
| Sex, n(%) | | | | | $\chi^2=4.63$ | 0.099 |
| Female | 139 (34.41) | 99 (34.02) | 31 (42.47) | 9 (22.50) |  |  |
| Male | 265 (65.59) | 192 (65.98) | 42 (57.53) | 31 (77.50) |  |  |
| T stage, n(%) | | | | | $\chi^2=8.58$ | 0.199 |
| T 1 | 32 (7.92) | 26 (8.93) | 4 (5.48) | 2 (5.00) |  |  |
| T 2 | 159 (39.36) | 120 (41.24) | 27 (36.99) | 12 (30.00) |  |  |
| T 3 | 138 (34.16) | 90 (30.93) | 27 (36.99) | 21 (52.50) |  |  |
| T 4 | 75 (18.56) | 55 (18.90) | 15 (20.55) | 5 (12.50) |  |  |
| N stage, n(%) |  |  |  |  | - | 0.086 |
| N0 | 15 (3.71) | 8 (2.75) | 3 (4.11) | 4 (10.00) |  |  |
| N1 | 38 (9.41) | 28 (9.62) | 3 (4.11) | 7 (17.50) |  |  |
| N2 | 255 (63.12) | 185 (63.57) | 49 (67.12) | 21 (52.50) |  |  |
| N3 | 96 (23.76) | 70 (24.05) | 18 (24.66) | 8 (20.00) |  |  |
| TNM, n(%) | | | | | $\chi^2=2.23$ | 0.327 |
| III | 247 (61.14) | 183 (62.89) | 39 (53.42) | 25 (62.50) |  |  |
| IVa | 157 (38.86) | 108 (37.11) | 34 (46.58) | 15 (37.50) |  |  |
| Differentiation degree, n(%) |  |  |  |  | - | 0.083 |
| WHO type I | 1 (0.25) | 1 (0.34) | 0 (0.00) | 0 (0.00) |  |  |
| WHO type II | 89 (22.03) | 71 (24.40) | 15 (20.55) | 3 (7.50) |  |  |
| WHO type III | 314 (77.72) | 219 (75.26) | 58 (79.45) | 37 (92.50) |  |  |
| EBV-DNA (copies/mL), n(%) |  |  |  |  | NA |  |
| ≤4 × 10<sup>2</sup> | 112 (27.72) | 89 (30.58) | 16 (21.92) | 7 (17.50) |  |  |
| >4 × 10<sup>2</sup> | 105 (25.99) | 73 (18.07) | 20 (27.40) | 12 (30.00) |  |  |
| Missing | 187 (46.29) | 129 (44.33) | 37 (50.68) | 21 (52.50) |  |  |

F: F-test, $\chi^2$ : Chi-square test, -: Fisher exact
SD: standard deviation
Abbreviations: WHO = world health organization, EBV-DNA = Epstein-Barr Virus DNA

### Prediction of Induction Chemotherapy Response

The predictive performance for IC treatment response is summarized in Table 2. TNM staging showed poor performance, with AUCs of 0.713 (95% CI: 0.620–0.806), 0.682 (95% CI: 0.596–0.768), and 0.645 (95% CI: 0.533–0.767) in the train, validation, and test sets, respectively. The unimodal DLR and Pathomics models demonstrated moderate performance. Specifically, the DLR model achieved AUCs of 0.774 (95% CI: 0.682–0.867), 0.711 (95% CI: 0.624–0.797), and 0.709 (95% CI: 0.587–0.831) in the train, validation, and test sets, respectively. The Pathomics model yielded AUCs of 0.832 (95% CI: 0.745–0.918), 0.772 (95% CI: 0.689–0.858), and 0.752 (95% CI: 0.680–0.824) across the same sets. In contrast, the integrated MoEMIL model attained the highest AUC in all datasets: 0.917 (95% CI: 0.848–0.986), 0.869 (95% CI: 0.790–0.947), and 0.801 (95% CI: 0.726–0.876) in the train, validation, and test sets, respectively, demonstrating a 22.3% improvement over the TNM staging and 15.6 % over DLR model, and a 9.3 % marginal improvement over the Pathomics model (Fig 3).

**Table 2.**
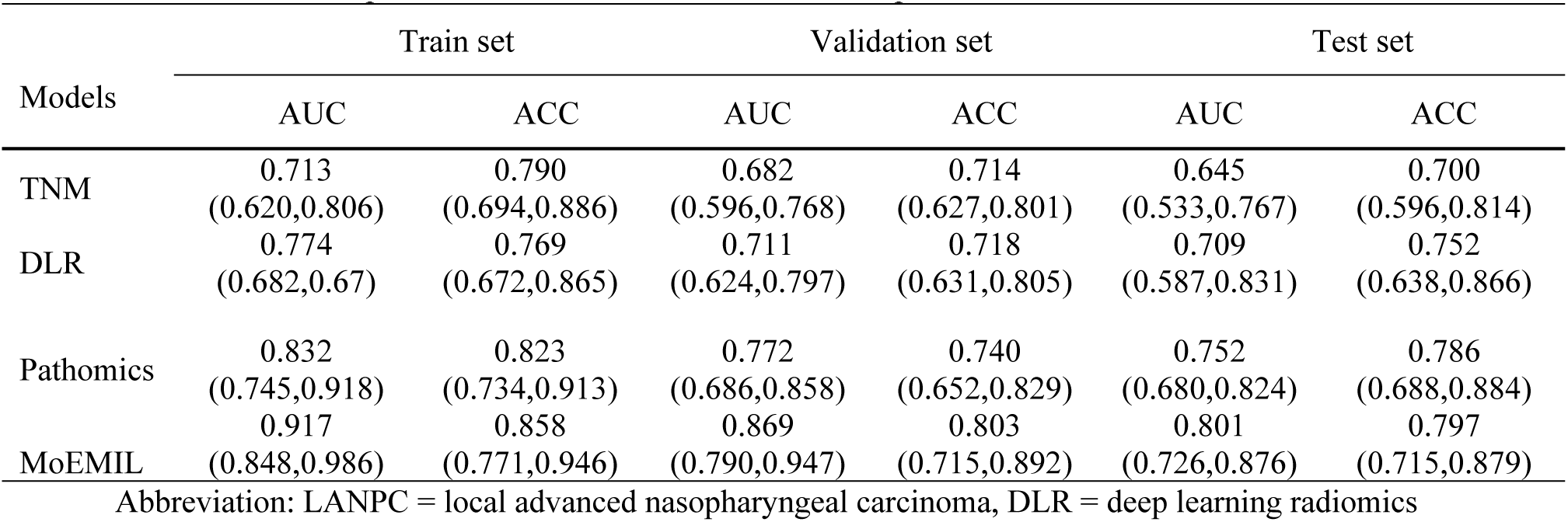
Predictive performance of three models in LANPC patients.

| Models | Train set |  | Validation set |  | Test set |  |
| --- | --- | --- | --- | --- | --- | --- |
|  | AUC | ACC | AUC | ACC | AUC | ACC |
| TNM | 0.713<br>(0.620,0.806) | 0.790<br>(0.694,0.886) | 0.682<br>(0.596,0.768) | 0.714<br>(0.627,0.801) | 0.645<br>(0.533,0.767) | 0.700<br>(0.596,0.814) |
| DLR | 0.774<br>(0.682,0.867) | 0.769<br>(0.672,0.865) | 0.711<br>(0.624,0.797) | 0.718<br>(0.631,0.805) | 0.709<br>(0.587,0.831) | 0.752<br>(0.638,0.866) |
| Pathomics | 0.832<br>(0.745,0.918) | 0.823<br>(0.734,0.913) | 0.772<br>(0.686,0.858) | 0.740<br>(0.652,0.829) | 0.752<br>(0.680,0.824) | 0.786<br>(0.688,0.884) |
| MoEMIL | 0.917<br>(0.848,0.986) | 0.858<br>(0.771,0.946) | 0.869<br>(0.790,0.947) | 0.803<br>(0.715,0.892) | 0.801<br>(0.726,0.876) | 0.797<br>(0.715,0.879) |

Abbreviation: LANPC = local advanced nasopharyngeal carcinoma, DLR = deep learning radiomics

**Fig 3.**
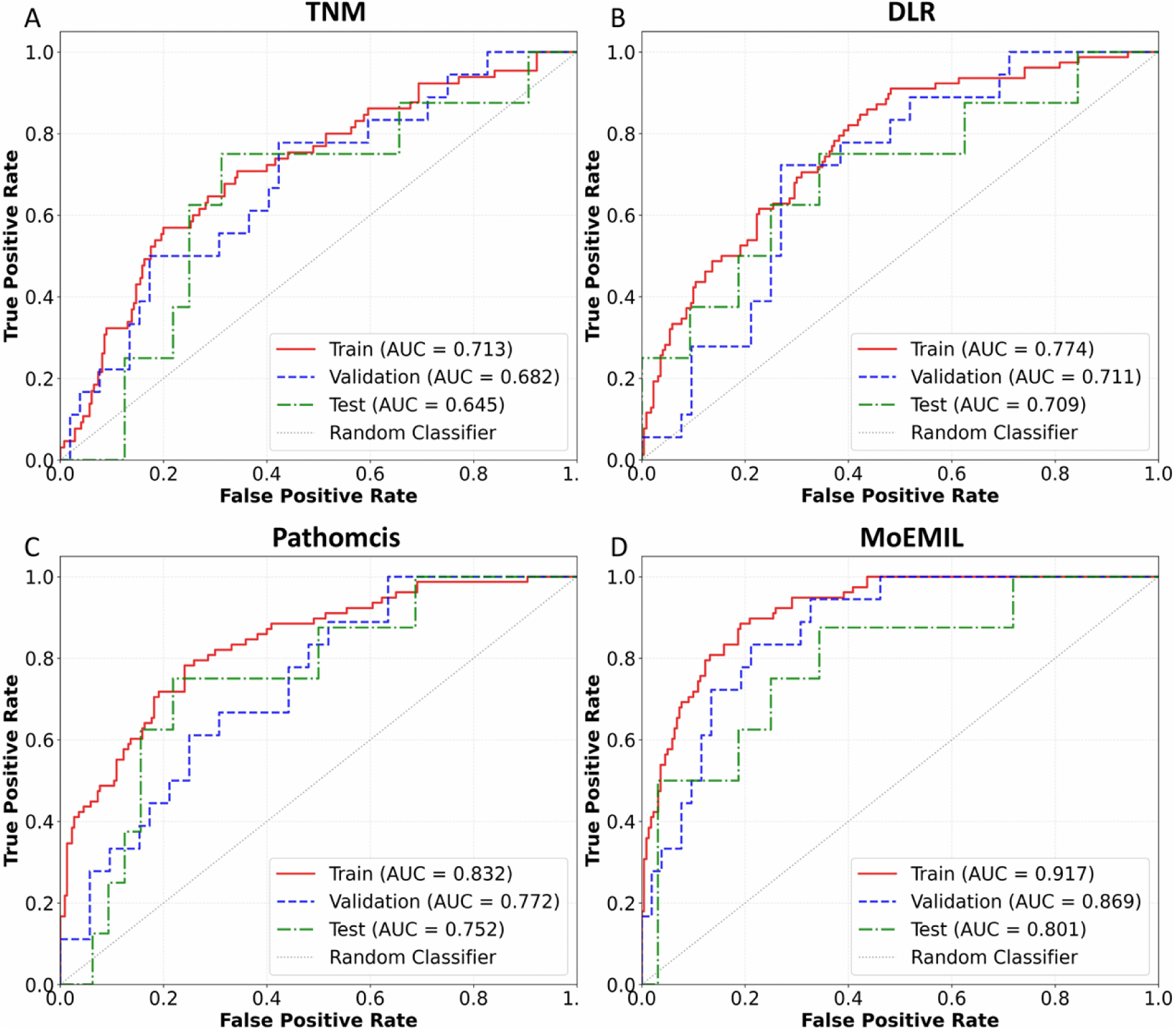
Predictive analysis for predicting IC response in LANPC patients using TNM staging, DLR, Pathomics, and MoEMIL models across both the training, validation, and external test sets. Abbreviations: OS = over survival, LANPC = local advanced nasopharyngeal carcinoma, DLR = deep learning radiomics

### Prognostic Performance for Overall Survival

The predictive performance for survival is summarized in Table 3. The TNM staging and DLR models showed relatively lower OS prognostic accuracy, with C-indices of 0.689(95% CI: 0.639–0.739) and 0.658 (95% CI: 0.605–0.710) in the train set, respectively. In contrast to DLR model, TNM staging demonstrated lower prognostic efficacy in both the validation and test sets (0.587 and 0.553 for TNM staging vs. 0.689 and 0.658 for the DLR model). In comparison, the Pathomics model exhibited improved performance, yielding C-indices of 0.765 (95% CI: 0.712–0.820), 0.681 (95% CI: 0.602–0.805), and 0.690 (95% CI: 0.650–0.701) across the same sets. The MoEMIL model outperformed both unimodal approaches and TNM staging, achieving the highest predictive accuracy across all cohorts: a C-index of 0.785 (95% CI: 0.701–0.854) in the train set, 0.773 (95% CI: 0.655–0.835) in the validation set, and 0.766 (95% CI: 0.701–0.853) in the test set. The DLR, Pathomics, and MoEMIL models successfully stratified patients into high- and low-risk groups, with Kaplan-Meier curves demonstrating statistically significant differences in OS (*P* < 0.05, Fig. 4). In contrast, TNM staging failed to achieve significant risk stratification in the training and validation sets (*P* > 0.05, Fig 4).

**Table 3.** Prognostic performance of three models in LANPC patients.

| Models | Train set |  | Validation set |  | Test set |  |
| --- | --- | --- | --- | --- | --- | --- |
|  | C-index(95%CI) | *P* | C-index(95%CI) | *P* | C-index(95%CI) | *P* |
| TNM | 0.689(0.639,0.739) | 0.012 | 0.587(0.537,0.637) | 0.014 | 0.553(0.463,0.643) | 0.041 |
| DLR | 0.657(0.605,0.710) | <0.001 | 0.655(0.603,0.707) | <0.001 | 0.676(0.564,0.767) | <0.001 |
| Pathomics | 0.765(0.712,0.820) | <0.001 | 0.681(0.602,0.805) | 0.009 | 0.690(0.650,0.701) | <0.001 |
| MoEMIL | 0.785(0.673,0.838) | 0.001 | 0.773(0.655,0.835) | <0.001 | 0.766(0.701,0.853) | <0.001 |

Abbreviation: LANPC = local advanced nasopharyngeal carcinoma, DLR = deep learning radiomics

**Fig 4.**
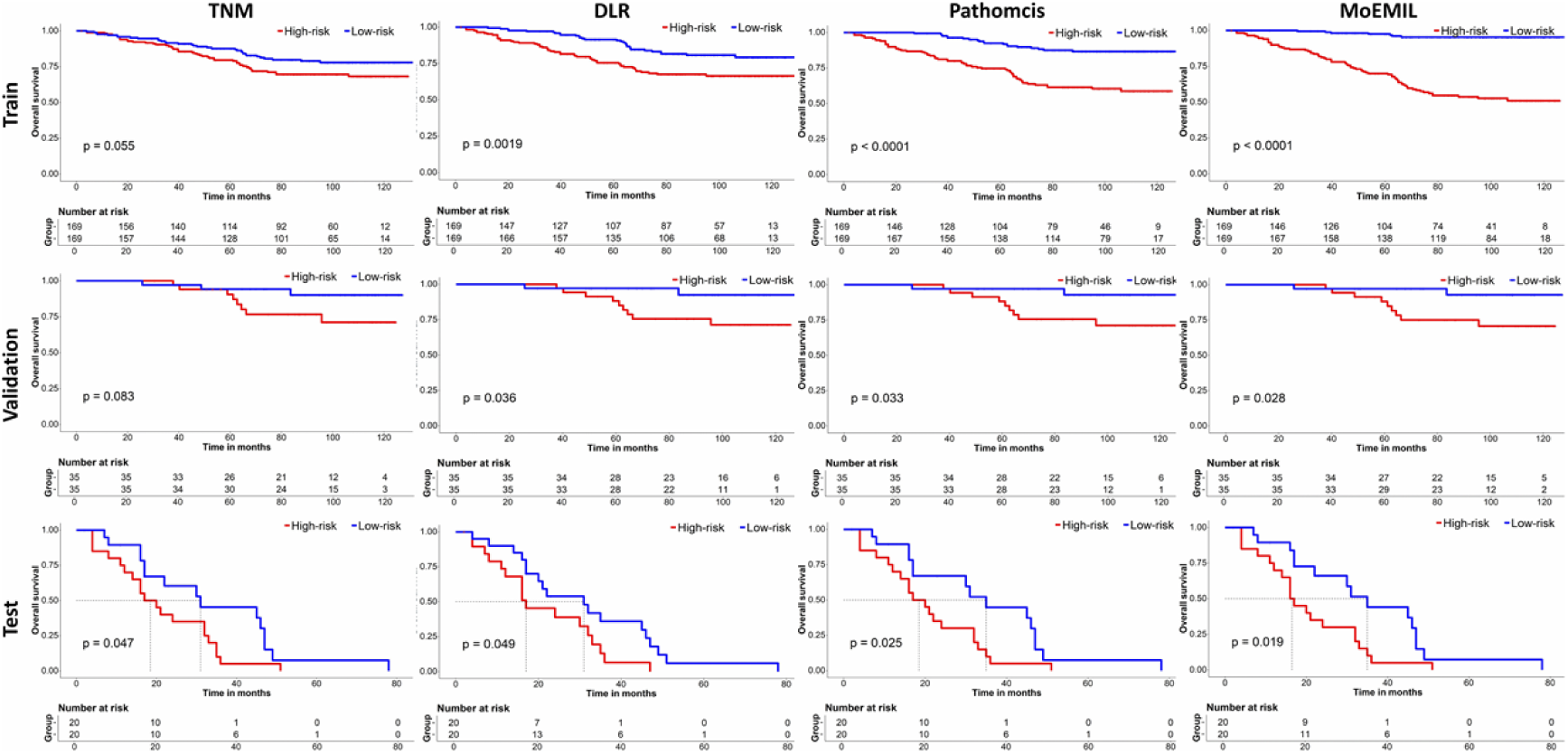
Risk stratification analysis for predicting OS in LANPC patients using TNM staging, DLR, pathomics, and MoEMIL models across both the train, validation, and test sets. Abbreviations: OS = over survival, LANPC = local advanced nasopharyngeal carcinoma, DLR = deep learning radiomics

### Interpretability and Visualization Analysis

For WSIs, we computed attention scores for all 256 × 256 non-overlapping tissue patches. These scores were then converted into percentile ranks ranging from 0.0 (low attention) to 1.0 (high attention) and spatially aligned with the corresponding WSI, averaging scores for any overlapping patches. The resulting heatmap was mapped to RGB values through a color map and overlaid on the original H&E stained image with 0.5 transparency. This high-resolution attention heatmap highlighted the relative prognostic relevance of different image regions used for risk prediction. We observed that high-attention regions in both high- and low-risk patients corresponded with tumor tissues (Fig 5A). However, for high-risk patients, high-attention regions in WSIs predominantly corresponded to tumor cell clusters which were large syncytiform cells with unclear cell boundaries, round or oval nuclei, and large centrally located nucleoli. For low-risk patients, high-attention regions in WSIs predominantly corresponded to tumor cell clusters that were fusiform and grew in clumps, with clear boundaries and keratinized cells occasionally. For MRI, the high-risk area was also highlighted on the tumor area based on Grad-CAM (Fig 5B), which highly overlapped with the lesion area identified by the WSIs. This further validates the robustness and clinical applicability of the model.

**Fig 5.**
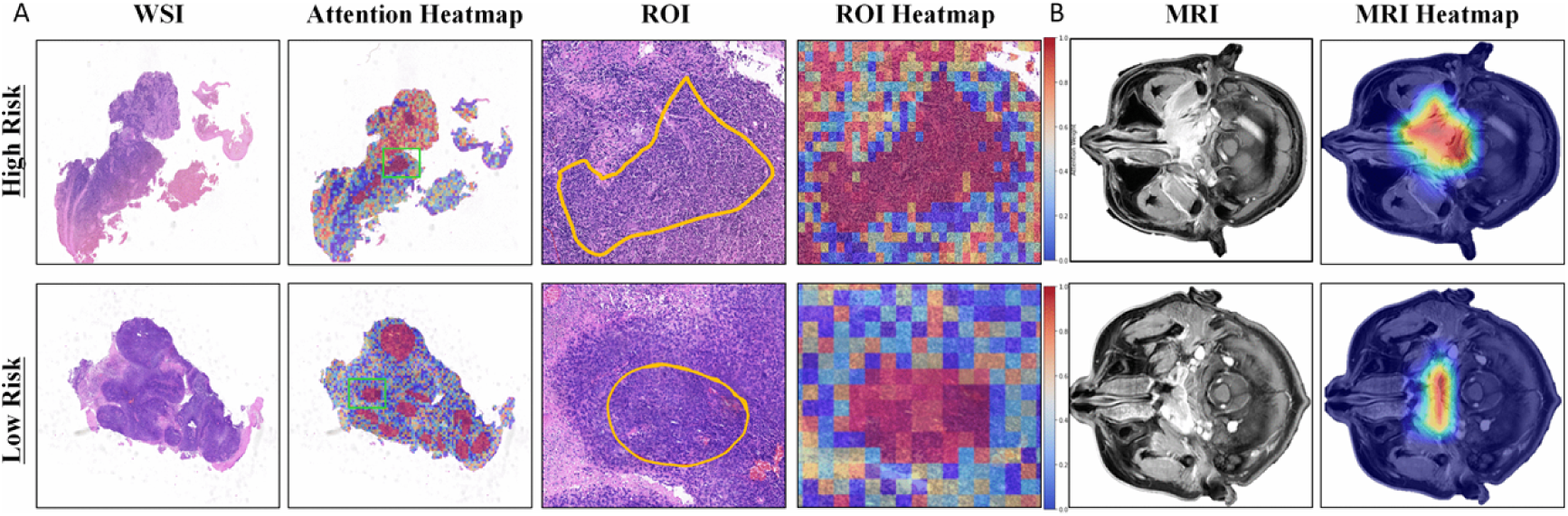
Interpretability and visualization. (A) Histological feature analysis: Sequential visualization from left to right includes the WSI thumbnail, attention-based risk heatmap, representative ROI, and the corresponding magnified ROI heatmap. For both high-risk (top row) and low-risk (bottom row) cohorts, the model’s attention predominantly aligns with viable tumor clusters and specific stromal microenvironments. The whole-slide attention heatmaps were generated by calculating MoEMIL-derived attention scores across overlapping patches (utilizing a 25% spatial overlap stride), providing a continuous landscape of the morphological features driving the risk prediction. (B) Radiological Explainability: Representative axial MRI scans displaying the primary tumor are paired with their corresponding Grad-CAM saliency maps. These heatmaps highlight the critical intra-tumoral regions that most significantly influence the model’s prognostic output. The Grad-CAM analysis confirms that the deep learning radiomics module effectively prioritizes the heterogenous core and invasive margins of the primary tumor, rather than background noise or non-specific features. Abbreviations: WSI = whole slide image, ROI = region of interest, Grad-CAM = Gradient-weighted Class Activation Mapping

## Discussion

In this multi-center study, a novel multi-task deep learning model integrating pretreatment MRI and WSIs was developed and validated to simultaneously predict IC response and survival outcomes in patients with LANPC. The proposed MoEMIL model demonstrated superior predictive performance compared to unimodal models in both internal and external validation cohorts. To our knowledge, this is the first study to integrate MRI and WSIs within a multi-task MMoE framework for dual-endpoint prediction in LANPC, extending prior MRI-only radiomics and deep learning approaches for prognostic evaluation in NPC[25] and for IC response and survival in LANPC[8,9].

Accurate pretreatment prediction of both IC response and survival outcome in LANPC patients is crucial for developing personalized treatment plans. Most existing models primarily depend on unimodal MRI data or integrate only clinical variables such as TNM staging and EBV-DNA status, neglecting the valuable insights that can be gained from multiscale image modalities. Radiomics captures tumor heterogeneity at the imaging level, reflecting macroscopic morphological and textural patterns, while pathomics, derived from WSIs, provides insights into cellular architecture and the tumor microenvironment. Our findings demonstrate that the DLR model relying solely on MRI data or the pathomics model relying solely on WSIs exhibits only limited to moderate performance in predicting IC response and survival outcomes. In contrast, the MoEMIL model, which integrates complementary macroscopic radiological and microscopic pathological information, substantially improves predictive efficacy. In both internal and external validation cohorts, the MoEMIL model consistently outperformed unimodal models in predicting IC response and survival outcomes. This finding aligns with the emerging paradigm of radiopathomics in other cancers^[16,18,26–30]^, reinforcing the principle that multi-scale data integration enhances prognostic modeling.

Notably, the proposed MoEMIL model demonstrated superior predictive performance for both IC response and OS compared to the conventional TNM staging system. While TNM staging remains the cornerstone of clinical decision-making in LANPC, it solely relies on anatomical extent and fails to capture the underlying tumor biological heterogeneity, which limits its ability to accurately stratify patients with divergent therapeutic outcomes. In our study, TNM staging exhibited limited predictive accuracy for IC response (AUC: 0.645–0.713) and poor prognostic discrimination for OS (C-index: 0.553–0.689), with Kaplan–Meier curves failing to show significant risk stratification in the training and validation sets. In contrast, the MoEMIL mode consistently achieved superior stratification across all cohorts. These findings underscore the inherent limitations of anatomical staging alone and highlight the added value of integrating multi-scale tumor characteristics through deep learning.

Multitask learning, as opposed to learning each task in isolation, is a joint learning paradigm that harnesses the inherent correlation of multiple related tasks to achieve reciprocal benefits in improving performance, enhancing generalizability, and reducing the overall computational cost[27]. In LANPC, tumor response to IC is an established independent prognostic factor for survival after IMRT[5]. Consequently, we hypothesized that IC response and OS share underlying biological mechanisms, such as tumor heterogeneity, cellular proliferation, and microenvironmental features, which manifest as common representations in high-dimensional feature space. By modeling these tasks jointly within a MMoE framework, our approach captures shared latent features through shared expert networks, while task-specific gating mechanisms dynamically filter information to account for the distinct nuances of short-term response versus long-term survival. This biological-to-computational synergy allows the model to learn more robust generalized features than single-task counterparts. The MoEMIL model effectively stratified patients into good- and poor-responders after IC, as well as stratified patients into high- and low-risk groups for OS. It achieved higher accuracy in predicting both IC response and OS compared to single-task models reported in the literature[8,9,28]. A key strength of the MoEMIL architecture lies in its dynamic feature-routing mechanism based on MMoE framework[17], where gating networks adaptively weight contributions from different experts according to each task-treatment response or survival prediction. This mechanism not only effectively utilizes shared information across multimodal data but also preserves task-specific features, thereby theoretically mitigating the common issue of negative transfer in multi-task learning and contributing to its consistent performance in external validation.

A further strength of this work is its emphasis on model interpretability and visualization. To elucidate the histopathological basis of predictions, we employed attention-based heatmaps that leverage learned attention scores to highlight tissue regions most influential to the model’s final outcome. In this study, high-attention regions in both high- and low-risk patients aligned with tumor areas, indicating that these regions significantly influence outcome prediction, which is consistent with our previous study[29]. In high-risk patients, these high-attention regions predominantly contained tumor cell clusters exhibiting large syncytiform cells with indistinct borders, round to oval nuclei, and prominent central nucleoli. Such morphology, as noted by Wang et al.[30], is associated with high proliferative activity, pleomorphism, and poor differentiation, features often linked to adverse prognosis. The heatmaps thus visually quantify each region’s contribution to risk prediction. Clinically, inspecting morphological details and other cellular features within high-attention areas on a WSI can aid in assessing the patient’s prognostic risk. To interpret the MRI model, Grad-CAM was applied for visualization. It calculates the gradients of the target class with respect to the final convolutional layer and uses global average pooling to produce localization maps, thereby identifying the most influential regions in the model ’ s decision-making process. Heatmaps intuitively display which regions of the input image are critical for the model’s prediction, helping understand the key areas the model focuses on. For instance, the WSI heatmap in Fig. 5A illustrates that the model focuses on tumor cell morphology, while the MRI Grad-CAM heatmap in Fig. 5B indicates the model’s attention to macroscopic tumor regions. Together, these two modalities contribute complementary information at different scales, collectively forming a comprehensive portrait of the tumor’s biological behavior. This phenomenon strongly suggests that macroscopic imaging representations and microscopic pathological morphology are not independent of each other, but rather are both rooted in the intrinsic, cross-scale biological behavior of tumors.

Our study has several limitations. Although this was a multi-center investigation, the sample size remains limited. Furthermore, as a retrospective study conducted over a long inclusion period, the model did not incorporate clinical factors-such as EBV-DNA, which was missing for a substantial proportion of patients-despite their established prognostic relevance in NPC. This reflects broader challenges in data completeness and consistency inherent to retrospective designs. As the first attempt to develop a MDL model for predicting IC response and OS in LANPC, its stability and generalizability require further validation with larger, multi-institutional datasets and prospective studies.

## Conclusion

In summary, we developed a novel multi-task deep learning model utilizing the MMoE framework to simultaneously predict IC response and OS by integrating radiological and pathological data. Further prospective multi-center studies are warranted to validate its clinical applicability in facilitating individualized therapy and prognostic decision-making.

## Data Availability

The original MRI and WSI data, as well as the clinical and follow-up data of the patients, are not publicly available due to restrictions imposed by the institutional ethics committees of Hunan Cancer Hospital and the First Affiliated Hospital of Jinan University, as well as patient privacy protection regulations under Chinese law. These data are available from the corresponding author upon reasonable request, subject to approval by the respective institutional data access committees and compliance with all applicable ethical and legal requirements. The radiomics features, deep learning features, and the histological patch?level features are publicly accessible without restriction. The code and trained model weights for the MoEMIL model are available at https://github.com/junjianli106/PathRadMoE.

## Acknowledgments

We would like to thank Storm Statistical Platform (www.medsta.cn) for statistical analysis.

## Notes

### Competing Interest Statement

The authors have declared no competing interest.

### Funding Statement

Yes

### Author Declarations

The study was approved by the Institutional Review Board of Hunan Cancer Hospital (Approval No.KY2026011).

